# Exclusive heat-not-burn cigarette smoking alters the profile of circulating microRNAs

**DOI:** 10.1101/2023.06.13.23291325

**Authors:** Vittorio Picchio, Giulio Ferrero, Claudia Cozzolino, Barbara Pardini, Erica Floris, Sonia Tarallo, Xhulio Dhori, Cristina Nocella, Lorenzo Loffredo, Giuseppe Biondi-Zoccai, Roberto Carnevale, Giacomo Frati, Isotta Chimenti, Francesca Pagano

**Author notes:** Equal contribution (Vittorio Picchio, Giulio Ferrero). Equal contribution (Isotta Chimenti, Francesca Pagano). Correspondence to: Francesca Pagano, Via E. Ramarini, 32 - 00015 Monterotondo Scalo, Rome, Italy.

## Abstract

**Background:** Traditional combustion cigarette (TCC) smoking is an established risk factor for several types of cancer and cardiovascular diseases. Circulating microRNAs (miRNAs) represent key molecules mediating pathogenetic mechanisms, and potential biomarkers for personalized risk assessment. TCC smoking globally changes the profile of circulating miRNAs. The use of heat-not-burn cigarettes (HNBCs) as alternative smoking devices is rising exponentially worldwide, and the circulating miRNA profiles in chronic HNBC smokers are unknown.

We aimed at defining the circulating miRNA profile of chronic exclusive HNBC smokers, and identifying potentially pathogenetic signatures.

**Methods:** Serum samples were obtained from 60 healthy young subjects, stratified in chronic HNBC smokers, TCC smokers, and Non Smokers (20 subjects each). Three pooled samples per group were used for small RNA sequencing, and the fourth sub-group constituted the validation set.

**Findings:** Differential expression analysis revealed 108 differentially expressed miRNAs; 72 exclusively in TCC, 10 exclusively in HNBC, and 26 in both smoker groups. KEGG pathway analysis on target genes of the commonly modulated miRNAs returned cancer and cardiovascular disease associated pathways. Stringent abundance and fold-change criteria nailed down our functional bioinformatic analyses to a network where miR-25-3p and miR-221-3p are main hubs.

**Interpretation:** Our results define for the first time the miRNA profile in the serum of exclusive chronic HNBC smokers and suggest a significant impact of HNBCs on circulating miRNAs.

**Funding:** Sapienza University of Rome grant 2019, # RG11916B8621D7FF to GBZ and grant 2021 # RM12117A5D7688BC to RC. IC is supported by grants # RM12117A805ED2FD and # RG11916B85CDBF76 from Sapienza University of Rome, and by grant # RG1221816BC8E766 from the Italian Health Ministry. FP is supported by Grant # A0375-2020-36621 from Regione Lazio (POR-FESR 2014-2021).

## Introduction

A healthy lifestyle is nowadays regarded as the best action for disease prevention to decrease both chronic and acute disease incidence in the general population. Among the modifiable risk factors, traditional combustion cigarette (TCC) tobacco smoking is the leading one against which clinicians fight, with a demonstrated causative role in the establishment and progression of cancer, lung diseases, chronic obstructive pulmonary disease, diabetes, and cardiovascular diseases (CVDs).^1–3^

Despite the efforts made worldwide in the last 15 years to reduce smoking through restrictions and higher taxation on tobacco, TCC use is still one of the main risk factors for premature mortality and morbidity, with the absolute number of smokers still showing an increasing trend in the world population.^4^

The mounting awareness of the harmful consequences of TCC smoking have pushed towards the development of alternative smoking devices, such as heat-not-burn cigarettes (HNBCs), which have been initially presented as safer than TCCs.^5^ However, HNBCs still have harmful consequences on health, despite at a lower extent, which qualifies them as modified risk products. This definition is mostly due to the lack of combustion-related specific toxic compounds. Moreover, the number of HNBC users is rising exponentially, already involving over 6% of the adult population in Europe, with alarming projections for the next decade.^6,7^

TCC smoking can harm every organ of the body, and its pathogenetic consequences have been long studied using model systems and in vitro assays on specific cell types. TCC smoke has been shown to affect the profile of circulating molecules detected in serum as a consequence of chronic, as well as occasional exposure to cigarette smoke. These molecules include reactive oxygen species (ROS), growth factors, and microRNAs (miRNAs) which can affect endothelial function, immune response, and other mechanisms which can trigger disease establishment in multiple organs.^8–13^

MiRNAs are a family of short single-stranded non-coding RNA molecules that regulate post-transcriptional gene silencing through base-pair binding on their target mRNAs^14^, thereby regulating virtually all cellular and biological processes.^15^

Circulating miRNAs have been shown to play an active part in mediating both physiological and pathological mechanisms.^16^ In addition, they can function as biomarkers for disease progression in cancer and CVDs, as well as for precision medicine applications and personalized risk assessment.^17,18^

Several transcriptomic studies have shown that TCC smoke exposure globally changes the profile of circulating miRNAs in both health and disease conditions.^12,19^ Both chronic and acute exposure to cigarette smoke can affect the presence of specific miRNAs in the bloodstream which can induce consequences in multiple cell types, including endothelial cells^11^, or lung cells.^13^ Changes in the circulating miRNAs can be reflected in specific organs, which show a modified miRNA expression profile after exposure to cigarette smoke.^13^

The effects of HNBCs on circulating miRNAs has not been thoroughly investigated yet. Given the relatively short time since their market launch, data on their long-term health effects (e.g., cancer, or chronic diseases incidence) are still unavailable. Some evidence on the effects on miRNAs has been collected in one animal study revealing a global downregulation of miRNAs in rats exposed for 90 days to mainstream smoke from a modified risk smoking device^20^ corresponding to the HNBCs used for our study.

In this paper, we aimed at profiling by next generation sequencing the circulating miRNAs in serum samples collected from young healthy individuals previously enrolled in the SUR-VAPES chronic clinical study, stratified based on chronic use of either TCCs or a specific HNBC device (mean exclusive use of HNBC=1.5±0.5 years), and compared them to matched Non Smoker subjects.^21^

## Methods

### Study approval

The samples used were collected for the SUR-VAPES chronic clinical study. Subject recruitment, participant characteristics, and experimental procedures were previously reported by Loffredo et al. ^21^ All the recruited subjects provided written informed consent to participate in the study which was granted approval by the Policlinico Umberto I Ethics Committee (protocol number 813/14) and conducted in compliance with the Declaration of Helsinki.

### Patients sample collection and experimental design

The serum samples had been previously collected from 60 healthy young subjects recruited for SURVAPES chronic study, stratified in three experimental groups of 20: chronic HNBC smokers (mean exclusive use of HNBC = 1.5±0.5 years), TCC smokers, and Non Smokers (NS) (M/F ratio and average age: 55% - 28 years in NS, 50% - 27 years in TCC, and 60% - 33 years in HNBC smokers). The sample size was estimated according to the measurements collected in the SURVAPES chronic trial, based on data from a previous study, considering a type I error probability α=0.05, power 1-β = 0.90, and a specific difference and standard deviation of the relevant parameters for the study^21^. The minimum estimated sample size for each group was n=17 which was increased to n=20.

Due to the lack of sufficient individual volumes of sera for a high-quality RNA extraction, we generated pooled samples following established guidelines in the literature.^22^

According to the sample size calculation method for RNAseq experiments ^23^, given a type I error α=0.05, a power 1-β = 0.80, and an effect size of 0.4 (applicable for human RNAseq datasets), the analysis of three samples at a sequencing depth of 10 million reads per sample would allow accurate detection of a log2 fold change of at least 1.68. Thus, we pooled our sera in order to have three samples for sequencing and, and focused only on miRNAs showing a log2FC of two for the functional analyses and validation.

The subjects were subdivided in four sub-groups of five each per experimental group using a simple randomization approach with the ‘split’ function in R. The anthropometric and available clinical parameters from the SUR-VAPES chronic study were tested to be homogeneous within each group and checked against the whole population, for each subgroup to be representative of the whole group of subjects. This was achieved through statistical analysis of both continuous and categorical variables (see paragraph Statistical Analysis). The sera of subjects in each subgroup were pooled in identical volumes to generate the samples for RNA extraction. Five serum samples per group were left un-pooled: RNA from these samples was extracted separately to constitute the validation set.

### RNA extraction and miRNAs quantification

The pooled serum samples were centrifuged at 16000rcf in order to remove debris, platelets or cells, and RNA was extracted from 200 µl, using the Qiagen miRNeasy kit (Qiagen GmbH, Hilden, Germany), following the manufacturer’s instructions. Syn-cel-miR-39 spike-in synthetic RNA (Qiagen GmbH, Hilden, Germany) was added to monitor extraction efficiency. RNA quality was assessed using Agilent 2100 Bioanalyser (Agilent Technologies Inc, Santa Clara, California, USA) and the Eukariote total RNA Pico kit (Agilent Technologies Inc, California, USA). RNA samples (6 µl for each pool) were used for library generation using the NEBNext Multiplex Small RNA Prep (New England Biolabs, Ipswich, Massachusetts, USA) as previously described by Galluzzo et al.,^24^, and sequenced on a single-read flow cell by bridge amplification and 75 cycles sequencing-by-synthesis on the Illumina NextSeq550 platform (Illumina Inc., San Diego, California, USA). The average sequencing depth obtained was 9.57 million +/- 2.15 raw reads.

The levels of miR-221-3p and miR-25-3p were quantified by absolute quantification using Qiagen LNA based SYBR green detection method (miRCURY LNA miRNA PCR assay-Qiagen). Briefly, 2 ul of total RNA was used to generate the cDNA according to the manufacturer protocol. The qPCR reaction was performed on the Applied Biosystems 7900HT machine, adding the relevant ROX concentration to the qPCR mix. For copy number determination, a standard curve was generated using cDNA derived from defined amounts of synthetic RNA for each miRNA. The miRNA amount was expressed as “copy number per well” of the qPCR plate as per the standard curve.

### Bioinformatic analysis

The small RNA-sequencing analyses were performed according to Tarallo et al..^25^ Briefly, reads were quality-controlled and trimmed of adapter sequences using Cutadapt v.3.7. Reads shorter than 14 nt were removed. Surviving reads were aligned on human miRBase v22.1 hairpin sequences using BWA v0.7.12. As described in Tarallo et al.,^25^ the pipeline quantifies mature miRNA sequences annotated in miRBase as well as “novel” mature miRNAs, identified based on the read mapping position within the hairpin sequence. Read count normalization and differential expression analysis was performed with DESeq2 v1.38.3. A miRNA was defined as Differentially Expressed (DE) if associated with a median number of reads greater than 15 in at least one study group and a Benjamini Hochberg (BH) adjusted p-value (adj. p) < 0.05.

Functional enrichment analysis was performed using RBiomirGS v0.2.12 considering only validated miRNA-targets interactions from miRTarBase v7.0 and miRecords. The analysis was performed with respect to MSigDB gene sets (libraries: c2.cp.kegg.v7.4, c2.cp.reactome.v7.4, and c5.go.bp.v7.4). Only terms associated with an adjusted p<0.05 in at least two gene targets were considered as significant. Semantic similarity analysis was performed using rrvgo v1.12 using the Resnik method and a similarity threshold of 0.85.

### Data availability

The raw and processed small RNA-Seq data were deposited in Gene Expression Omnibus with the identifier GSE233237.

### Statistical analysis

Samples were randomly assigned to each of the four subgroups, and statistical difference in each variable described was assessed by either Fisher’s exact test for categorical variables, or Wilcoxon rank sum test for continuous variables. Correlation analyses were performed using the Spearman’s method. These analyses were performed in R using the basic statistics package.^26^ Real-time PCR data were analysed using ordinary one-way ANOVA and Fisher LSD post-test on Prism8 Software (GraphPad Software, Boston, Massachusetts, USA).

## Results

### HNBCs change miRNA levels in substantial overlap with TCCs

Serum samples were collected from three groups of healthy young subjects, with homogeneous anthropometric features, and stratified according to smoking status: Non Smokers (NS), TCC smokers, and exclusive HNBC smokers (mean exclusive use = 1.5±0.5 years). All smokers were considered chronic as they had been using each device for at least one month. The abundance of miRNAs was detected in pooled sera samples, and data analyzed using DESeq2 (Figure 1a). The principal component analysis showed a neat separation between NS and both smoker groups, and a less pronounced difference between TCC and HNBC smoker sera (Figure 1b). Head-to-head comparison of the detected miRNAs showed that a higher number of DE miRNAs had changed levels in sera from TCC as compared to HNBC smokers (Figure 1c,d), thus suggesting a globally lower impact of HNBCs on the profile of circulating miRNAs in healthy subjects, compared to TCC use.

**Figure 1.**
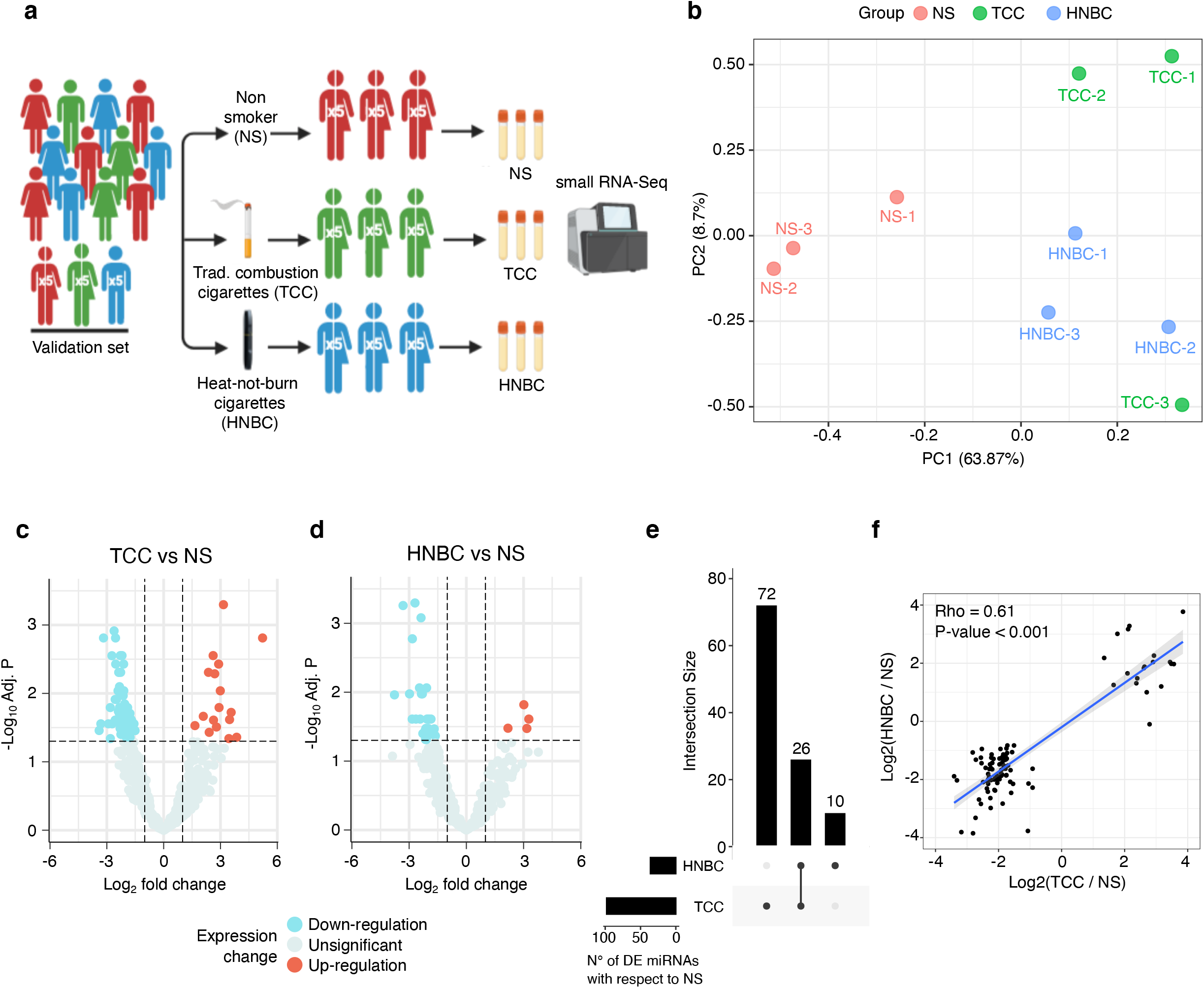
Analysis of circulating miRNAs in the serum of smokers. a) Experimental design of the study. Three pools of serum composed of five subjects each were generated starting from a sample population of 20 subjects. The remainder five samples were not pooled and used as single subjects for validation of differentially expressed miRNAs. b) Principal component analysis of the small RNAseq dataset. c-d) Volcano plots showing the Log2 Fold Change vs –log10 p value of the differentially expressed miRNAs (DE miRNAs) in each group of smokers compared to Non Smokers. Dashed lines indicate the log2FC threshold set to 1 (vertical) and the –log10P threshold, set to a value corresponding to the –log10(0.05) on the linear scale. e) Upset plot showing the number of Differentially Expressed (DE) miRNAs in each group and the intersection, which constitutes the class of common DEmiRNAs. f) Correlation plot of the Log2FC values in each comparison, showing the linear correlation of the changes observed in each group of smokers compared to the Non Smokers.

Differential expression analysis identified 98 differentially expressed (DE) miRNAs in TCC and 36 DE miRNAs in HNBC smokers, with an overlap for 26 miRNAs (Supplementary Table1, and Figure 1e). The comparison of the fold change of DE miRNAs in TCC and HNBC versus NS control shows a positive correlation (rho=0.61, p<0.001), which indicates that exposure to TCC or HNBC smoke implicates changes in the circulating miRNA levels in the same direction, but possibly to different extents (Figure 1f).

The profile of DE miRNAs revealed an overall downregulation of circulating miRNA abundance, with only 20 miRNAs up-regulated in both groups of smokers versus NS (Figure 2a), consistently with previous studies.^19,27,28^ Clustering analyses showed the similarity between the TCC and HNBC groups, with extensive overlap in the list of miRNAs commonly affected by both smoking types (Figure 2a). The detected level of DE miRNAs in HNBCs as compared to NS, tends to an intermediate level between the NS and the TCC groups for both upregulated and downregulated miRNAs (Figure 2b).

**Figure 2.**
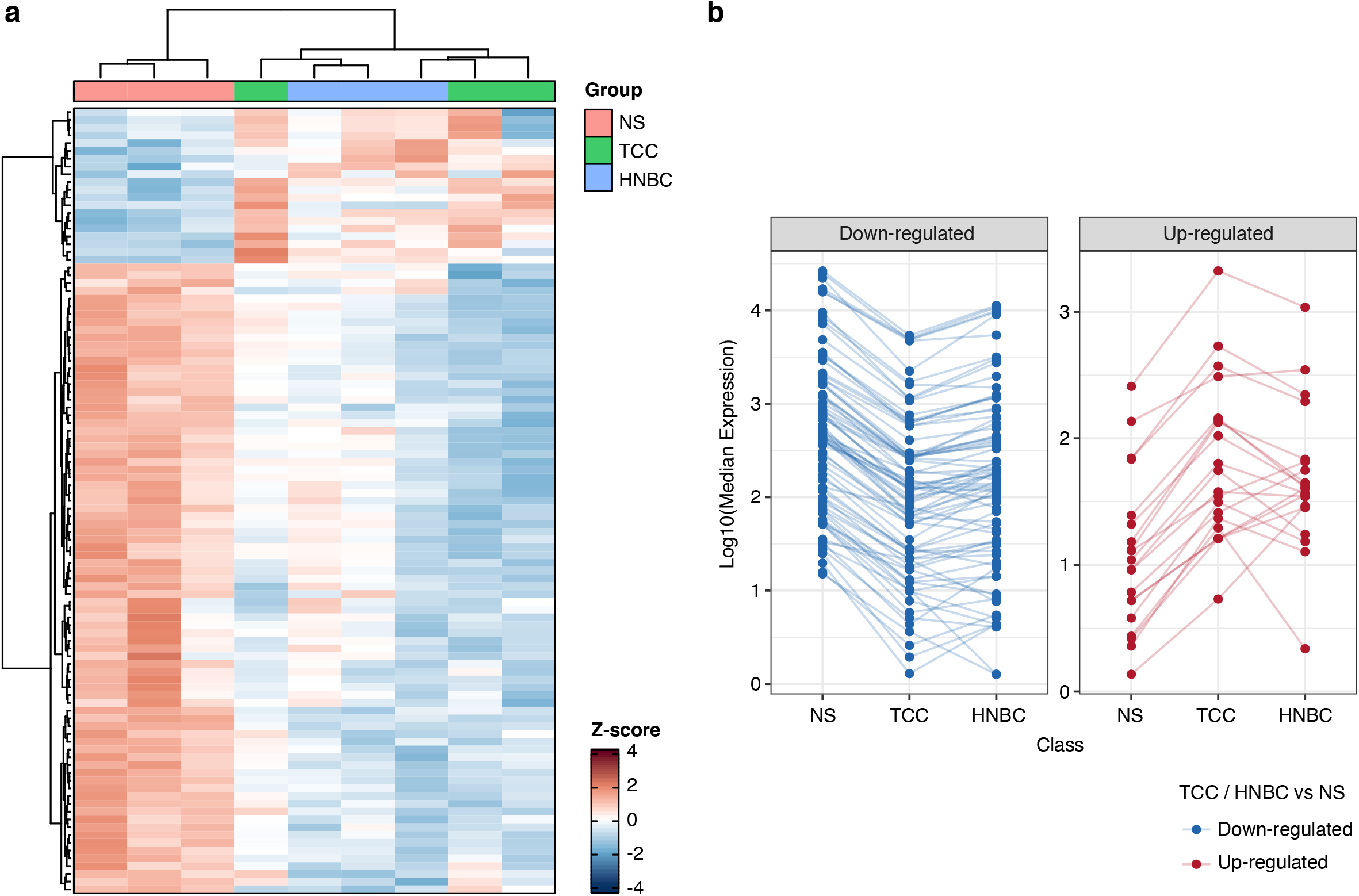
Clustering and global level of differentially expressed circulating miRNAs in TCC and HNBC smokers. a) Heatmap with hierarchical clustering showing the differentially expressed miRNA clustering in sera of Non Smokers (NS), Traditonal Combustion Cigarette (TCC) and Heat Not Burn Cigarette (HNBC) smokers. b) Line plot showing each single detected miRNA abundance in the three groups (Median expression value) for both downregulated and upregulated miRNAs.

### Functional analysis of commonly deregulated miRNA targets points to cancer and CVD

To focus the analysis on a subset of potentially functional circulating molecules, a list of highly abundant DE miRNAs (median normalized count greater than of 500 reads in at least one group) was isolated, which included the top 25% of the DE miRNAs in the analysis (Table 1). Based on the comparison in which the miRNA was identified as significantly DE, the list was divided in three classes: TCC-specific (TCC-var), HNBC-specific (HNBC-var), or common to both categories (COMMON). The clustering of samples using these miRNAs recapitulated that obtained with the whole DE miRNA list (Figure 3a). Of note, the HNBC-var class contained only one miRNA (miR-941), thus the effect of HNBC on circulating miRNAs in our datasets is almost completely included in that of TCC. All miRNAs included in these lists were downregulated, except for miR-200a-3p and miR-200b-3p, which were upregulated in the TCC group (Figure 3a).

**Table 1:**
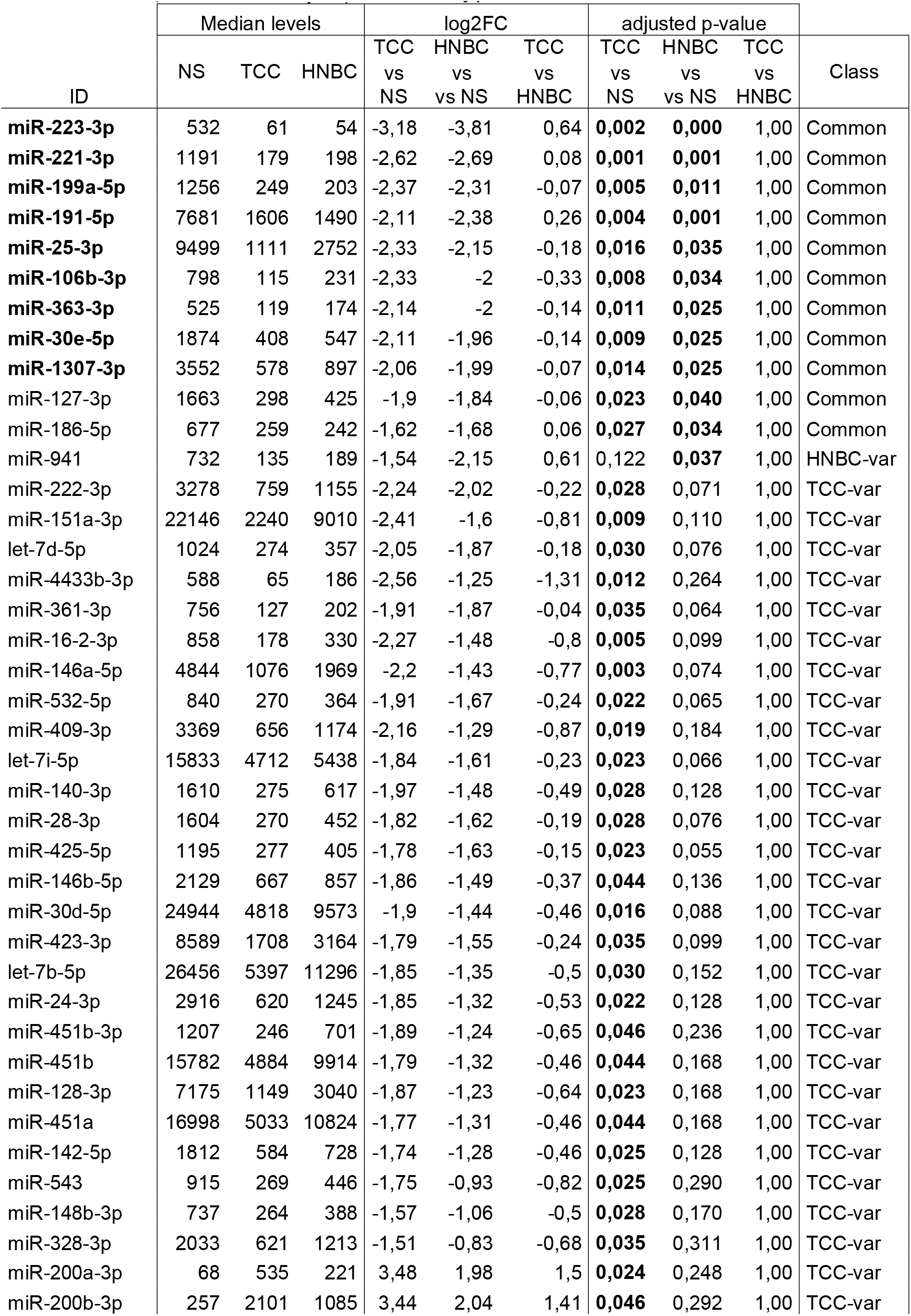
Selection of differentially expressed microRNAs. Table showing the list of miRNAs selected according to abundance in serum (counts>500). Bold character in the “adjusted p-value” column shows the significant p values. Bold character in the ID column shows the differentially expressed miRNAs with a fold change greater than two in the Common class.

**Figure 3.**
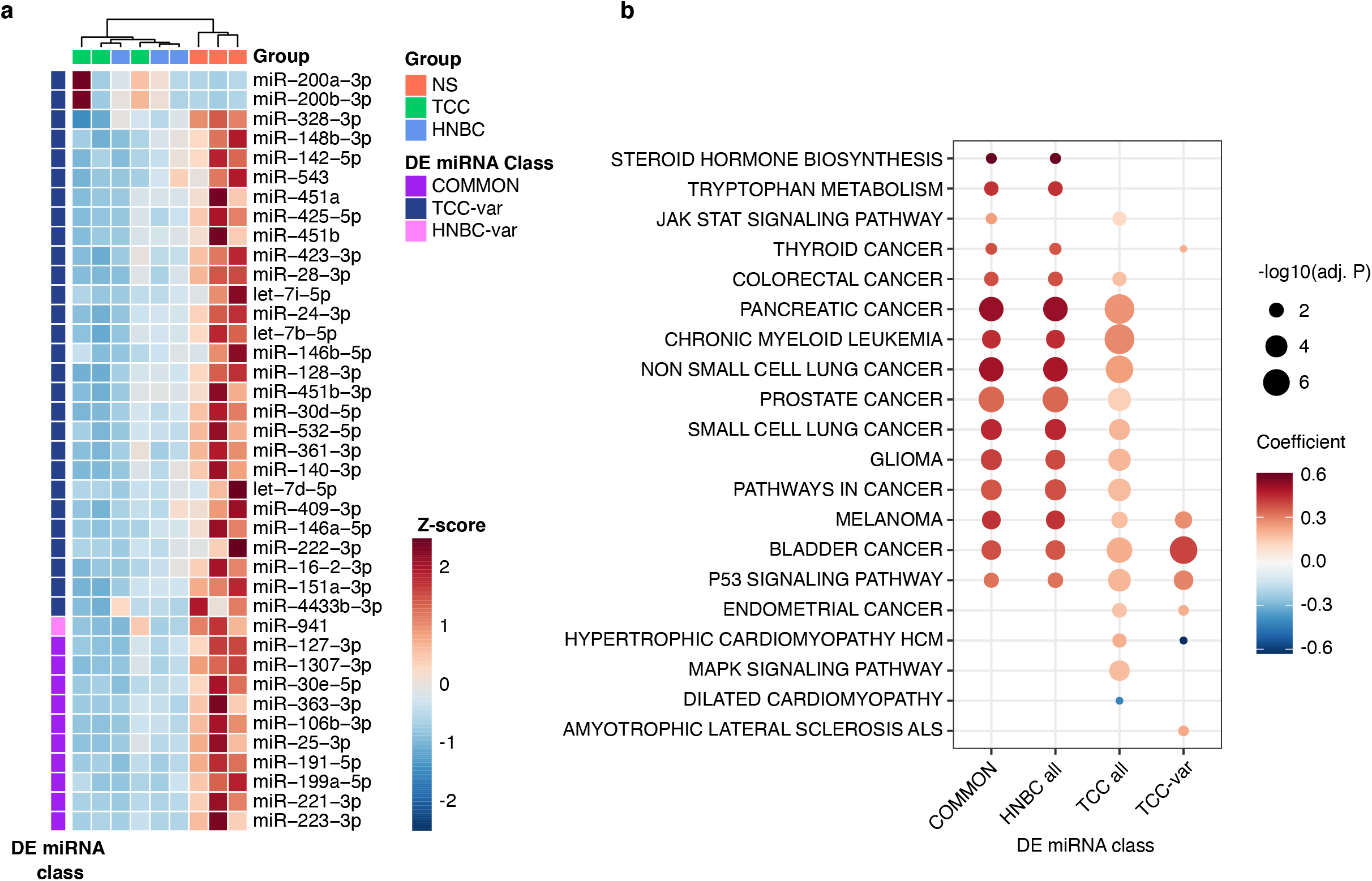
Highly abundant circulating miRNAs and the functional pathways affected. a) Heatmap with hierarchical clustering showing the relative fold-change of the sub-list of differentially expressed miRNAs (DE miRNAs) selected according to abundance in serum (counts>500). The code colour in the legend shows the sample groups, as well as the classification according to whether the miRNA level changes are detected in either both types of smokers (Common) or only in one of the two (TCC_var and HNBC_var). b) Bubble graph showing the enriched terms in KEGG functional analysis of the targets of the DEmiRNAs changing in both groups (Common), in the HNBC group including the class Common (HNBC all), in the TCC group including the class Common (TCC all), and only in TCC group (TCC-var). Bubble size is linearly correlated with the significance of the enrichment (-log10 adjp. value) and bubble colour is related to the coefficient, which is the predicted up- (red) or down-regulation (blue) of each listed pathway. TCC = Traditional Combustion Cigarette, HNBC = Heat Not Burn Cigarette, NS = Non Smokers.

The global downregulation observed suggests a de-repression of molecular pathways regulated by these miRNAs. The functional analysis was performed on the target genes of the DE miRNAs commonly deregulated in TCC and HNBC, only in one smoking group, and on a fourth group containing the whole list of TCC deregulated miRNAs (TCC-only = COMMON + TCC-var), to have a reference for the pathways potentially affected by TCC on its own. The Gene Ontology (GO) analysis highlighted 733 enriched biological processes that were clustered to 191 parent terms based on their semantic similarity (Supplementary Table 2-3). It also showed that the most significantly enriched terms refer to miRNAs deregulated in the COMMON class, which almost completely overlaps with the HNBC-var class (Supplementary Figure S1), suggesting that the effects of both kinds of smoking on circulating miRNAs affect similar biological processes. The GO terms enriched in the targets of miRNAs in the COMMON category refer to cell differentiation, cell cycle control, and immune response (Supplementary Figure S1).

The molecular pathways regulated by the affected miRNAs were further investigated using a KEGG analysis approach, which retrieved a relevant number of cancer-associated pathways enriched in targets of DE miRNAs, followed by signalling and CVD pathways (Figure 3b, Supplementary Table 2). The comparison of KEGG terms between the TCC-only and COMMON/HNBC-var categories showed that the terms in the latter group are still enriched in cancer-related pathways. This observation points to a deregulation of circulating miRNAs in chronic HNBC smokers targeting pathways known to be altered in cancer and overlapping with the deregulation observed in TCC smokers. The KEGG analysis was in line with the GO analysis where the enriched terms were related to neoplastic transformation of cells and CVD emergence (Supplementary Figure S1).

A miRNA-target network was finally defined to identify genes possibly affected by the observed miRNA deregulation in blood and target organs. The analysis was limited to miRNAs with at least 500 normalized counts and decreasing at least two-fold in the COMMON category, thus restricting to 10% of all the DE miRNAs in the dataset (Table 1 and Supplementary Figure S2). Then, the network was designed by including only miRNA-gene interactions experimentally validated (Supplementary Table 4). This set of stringent parameters identified a core network composed of nine miRNAs and 203 genes organized in two main hubs (Figure 4a). In this network, the genes targeted by the most-connected miRNAs (i.e. miR-25-3p and miR-221-3p) are involved in cancer and CVD-related pathways. Specifically, miR-25-3p was connected with cancer- and CVD-related genes, while miR-221-3p prevalently with cancer-related genes (Figure 4a). The expression of these two miRNA hubs was tested in the independent validation set of five subjects per group through quantitative real-time PCR, and data confirmed the expression levels observed in the small RNA-Seq dataset (Figure 4b).

**Figure 4.**
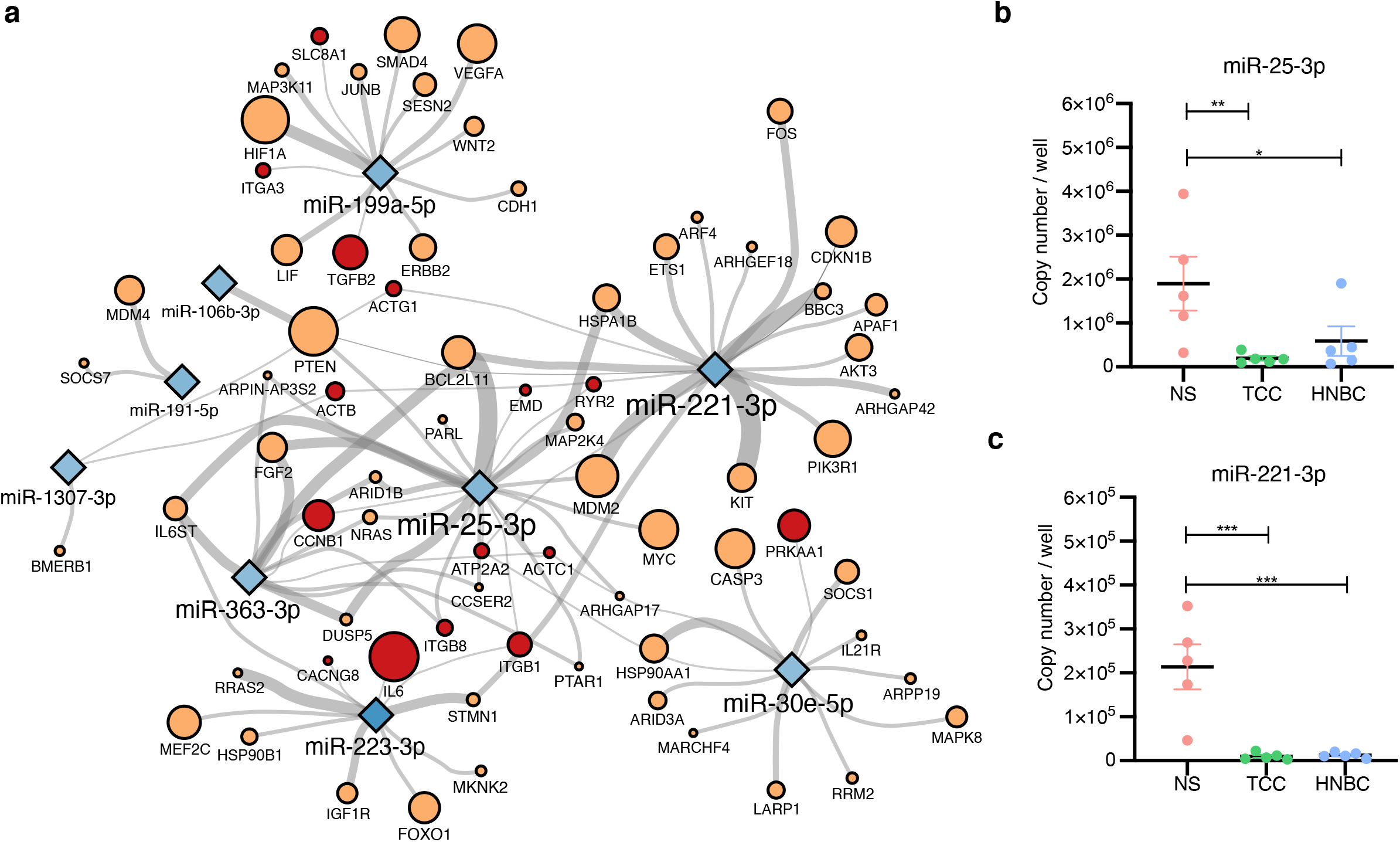
Network for target analysis of selected DEmiRNAs. a) Network of the miR/mRNA connections of the most abundant and highly changing miRNAs in both TCC and HNBC smokers (counts>500 and fold-change >2) compared to NS. The size of each gene is proportional to the number of times the term occurs within the enriched pathways in our functional analysis. The edges thickness is proportional to the number of evidence supporting the miR-mRNA interaction. The colouring of genes refers to the gene being present in Cardiovascular disease- (red) or Cancer- (orange) related pathways. b) Dot-plot showing the copy number obtained by realtime qPCR absolute quantification of the two hub miRNAs identified, i.e. miR-25-3p and miR-221-3p. Each dot shows the copy number/well in the PCR reaction as obtained by the standard curve method. Error bars show S.E.M. Statistical analysis was performed using Two-way ANOVA with multiple comparison testing. *= p<0.05, **= p< 0.01 *** p< 0.001. TCC = Traditional Combustion Cigarette, HNBC = Heat Not Burn Cigarette, NS = Non Smokers.

## Discussion

The use of extracellular miRNAs as biomarkers has been proven effective as well as minimally invasive for the detection and follow-up of many diseases. Indeed, changes in circulating miRNAs have been associated with many pathological conditions, such as (but not limited to) type 2 diabetes, obesity, CVD, and cancer. Therefore, alterations occurring in healthy individuals due to specific habits such as smoking, should be well described and considered when assessing the reliability of specific miRNAs as disease biomarkers. The miRNAs identified here as correlated with the chronic use of HNBC may be biomarkers of exposure to this specific alternative smoking device, and may regulate gene expression in target organs.

The identification of a signature of miRNAs altered by chronic smoking of both TCC and HNBC in healthy subjects is relevant in the use of miRNAs as disease biomarkers. For example, it has been reported that TCC smoking can affect the reliable identification of circulating miRNAs, including miR-25-3p, as diagnostic biomarkers in lung cancer^29^, hence the smoking status, including the use of modified risk products such as HNBCs, might interfere with the monitoring of the disease. The analysis of circulating miRNAs may also serve as an indicator of the physiological status of the subjects, and changes in the circulating miRNAs have been correlated with differences in lifestyle activities, such as exercise or smoking, in otherwise healthy people^19,30^, as well as in physiological aging^31^. In this unique and homogeneous group of healthy young individuals smoking exclusively either TCCs or HNBCs, enrolled in the SUR-VAPES chronic trial, the effects of TCC smoke on circulating miRNAs seem to be extensively recapitulated by the modified risk product namely HNBCs. The changes detected in circulating miRNAs of young healthy smokers using TCC as well as HNBC exclusively, might predispose these subjects to the development of diseases, or to have a weaker response to organ injury or damage^32^.

Interestingly, we observed a common trend for the abundance of circulating miRNAs modulated in HNBC smokers to be positioned at intermediate values between the NS and the TCC smokers. Despite with some limitations due to the pooling of samples and the lack of a matched control group of former smokers (many of the HNBC smokers in our study are in fact former TCC smokers, with abstinence from TCC since at least 1.5 years^21^) our data cannot rule out a partial contribution of a wash-out effect in the HNBC group. Nonetheless, it has been reported that quitting smoking brings the profile of most plasma miRNAs back to levels comparable to Non Smokers in one month^19^, and the subjects enrolled in the present study in the HNBC group have been exclusively smoking HNBCs for about 1.5 years on average. Therefore, our data can indeed point out that some fundamental alterations on circulating miRNAs are maintained when switching from TCC to chronic exclusive HNBC smoke, albeit to a lower global extent compared to chronic TCC use.

Many GO terms and pathways identified in our study from the list of altered miRNAs are linked with leukocyte biology and differentiation. One might speculate that potential systemic effects of the observed circulating miRNA depletion might contribute to disease development also via altered inflammatory responses^33^. Our final stringent network analysis evidenced two main hubs, with interesting interactions and targets from a pathological perspective. The precursor of miR-25-3p is part of a genomic cluster, highly conserved in vertebrates, which includes miR-106b and miR-93 pre-miRNA. Three mature miRNAs of this cluster show downregulation in the serum of smokers, with miR-25-3p being the most abundant among the three members of the cluster (9499 normalized reads for miR-25-3p, compared to 396 of miR-93-5p, and 797 of miR-106b-3p, as per the normalized expression values in DE analysis, in the NS group). Deranged expression of miR-25-3p has been associated with multiple cardiovascular and respiratory insults and diseases. This miRNA was shown to be regulated in a time-dependent way in cardiac hypertrophy, fibrosis, and heart failure^34^. In many systems and after different pathological stimuli, miR-25-3p decrease determines an increase in oxidative stress. This effect is mediated by the de-repression of *NOX4*, as demonstrated in cardiac injury, as well as in a kidney model of streptozotocin-induced type II diabetes^35,36^. This is also consistent with the general increase in ROS and oxidative stress markers previously described in our samples in the SUR-VAPES chronic study^21^. Thus, a lower level of circulating miR-25-3p could predispose the organism to oxidative stress in multiple organs. Moreover, the miR-25 cluster has shown protective effects against TGF-beta-induced epithelial-to-mesenchymal transition, which is responsible for tissue fibrosis in different organs, including the heart^37–39^. MiR-25-3p has also a protective role against apoptosis in both heart and brain ischemia/reperfusion injury, as demonstrated in rat ventricular myocytes and in neuronal cell models^40,41^. Finally, atherosclerosis development with intimal hyperplasia was also linked to miR-25-3p repression in vascular smooth muscle cells subjected to thrombospondin treatment^42^. In addition to CVD, miR-25-3p was demonstrated to be repressed in human tracheal smooth muscle cells exposed to pro-inflammatory cytokines, as a model of bronchial asthma. The repression of miR-25-3p resulted in the increase of its target gene Krüppel-like factor 4 (*KLF4*), a well-known mediator of inflammation^43^. Tobacco smoking from both TCC and HNBC is associated with asthma symptoms and with a faster development of lung disfunction^44^. Overall, miR-25-3p could mediate disease mechanisms associated with smoke-induced increase in airway epithelia inflammation.

MiR-221-3p is the second main hub identified by our network analysis (Figure 4a), and it has been described as both an oncogene and a tumour suppressor, depending on the cellular context. In particular, miR-221-3p was found to: i) inhibit cell proliferation and induce apoptosis in various cancers, such as erythroleukemia and gastrointestinal stromal tumour pathogenesis^45,46^; ii) inhibit the proliferation and migration of epithelial ovarian cancer cells through downregulation of *ARF4*^47^; and iii) to promote cell cycle arrest and apoptosis, and inhibit proliferation in medulloblastoma cells^48^. Therefore, reduced miR-221-3p levels point to several tumour-promoting pathways. The circulating levels of miR-221-3p are also downregulated in atherosclerosis and stroke, where they can be used as a biomarker^49,50^. Interestingly, the molecular mechanism whereby low levels of miR-221-3p might accelerate atherosclerosis has been linked with oxidative stress increase. In fact, miR-221-3p was found to inhibit ox-LDL-induced foam cell formation, to suppress -LDL-induced oxidative stress and apoptosis in monocytes, thus opposing to atherosclerotic plaque formation and progression. Notably, the subjects included in the present study have been previously assessed for vascular function, oxidative stress, and platelet activation, all of which were found impaired in the HNBC cohort of the SURVAPES chronic study^21^. The increase in oxidative stress and platelet activation was assessed through the measurement of different soluble markers; interestingly, the levels of H_2_O_2_, sNOX2-dp, and sCD40L were inversely correlated with those of circulating miR-221-3p, with sCD40L also correlating with miR-25-3p levels, in the 15 individual samples from the validation set (Supplementary Figure S3). It is worth noticing that both miRNAs significantly correlated to the cotinine levels in this same validation set.

We acknowledge several limitations of our study. The sample pooling has not allowed correlation analyses on available blood parameters or vascular function for all the subjects, and it has not allowed a thorough assessment of the validity of the detected DE miRNAs as markers for exposure to smoke from TCCs or HNBC devices. Also, working with human samples from healthy subjects, we could not provide insights on the correlation between circulating miRNA levels and their expression in organs and tissues. Nevertheless, to the best of our knowledge the data collected here are the first evidence of the effects of chronic exclusive HNBC smoking on the circulating miRNAs in young healthy individuals.

In conclusion, our findings will need to be further developed in future studies to provide insights on the efficacy of circulating miRNAs as biomarkers for early-stage diseases linked with smoke exposure from both sources, as well as on the molecular mechanisms leading to cellular dysfunction through impaired miRNAs signalling due to smoke exposure.

## Data Availability

All data produced in the present work are either contained in the manuscript or available at GEO.

https://www.ncbi.nlm.nih.gov/geo/query/acc.cgi?acc=GSE233237

## Acknowledgements

This study was partially funded by Sapienza University of Rome, grant 2019, Prot. RG11916B8621D7FF to GBZ, grant 2021 # RM12117A5D7688BC to RC. IC is supported by grants # RM12117A805ED2FD and # RG11916B85CDBF76 from Sapienza University of Rome, and by grant # RG1221816BC8E766 from the Italian Health Ministry. FP is supported by Grant # A0375-2020-36621 from Regione Lazio (POR-FESR 2014-2021).

## Contributors

VP, FP conducted experiments and analysed data; GFe designed and performed bioinformatic analyses; BP, ST conducted experiments and provided reagents; CC, EF conducted experiments; XD analysed data; CN, LL, RC, GFr, acquired clinical data and provided the serum samples; GBZ statistical analysis; IC, FP designed the study and wrote the manuscript.

