## Supplementary figures for "Exclusive heat-not-burn cigarette smoking alters the profile of circulating microRNAs"

### Supplementary material

#### Table of contents

Supplementary Figure S1
Supplementary Figure S2
Supplementary Figure S3

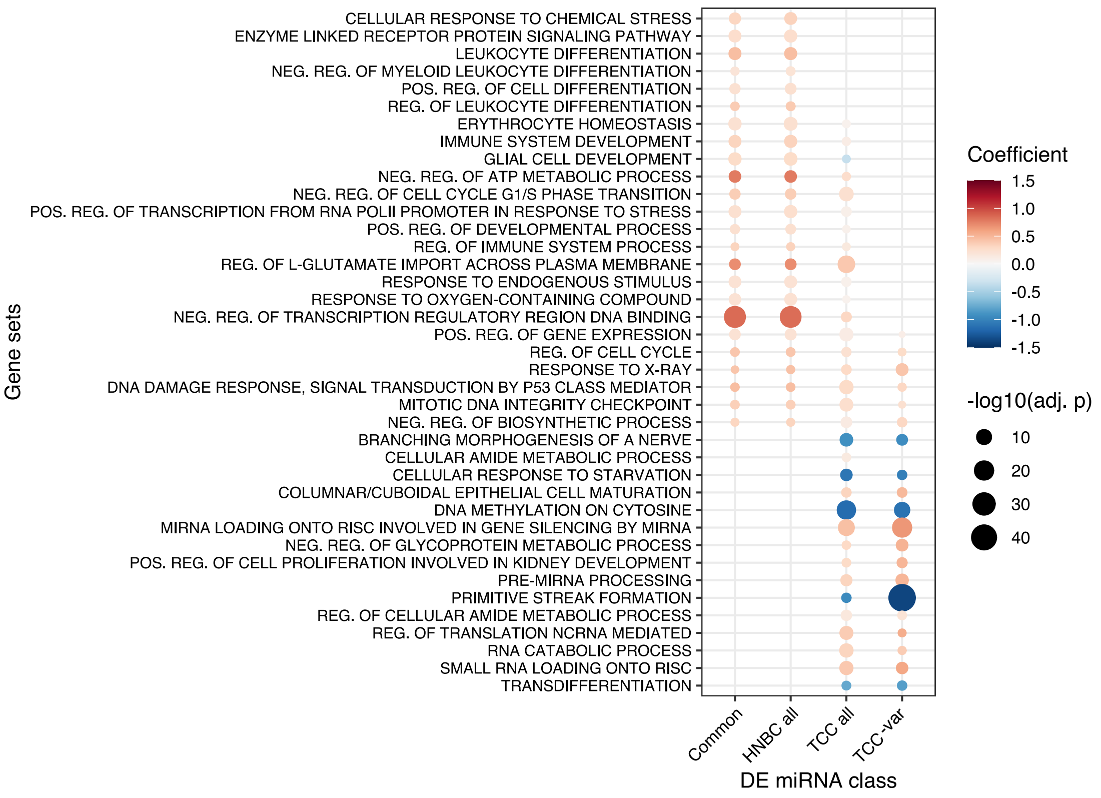


**Supplementary Figure S1**

**Gene ontology analysis for the target genes of the differentially expressed miRNAs**. Bubble plot showing the enriched terms in the GO Biological Process category obtained by the analysis of validated target genes of the differentially expressed miRNAs. The size of the bubbles is proportional to the significance of the enrichment. The colour code refers to the coefficient computed by RBiomirGS. Negative (in blue) and positive (in red) coefficients represent processes predicted to be down-regulated or up-regulated respectively, based on the DEmiRNA expression change in each comparison.

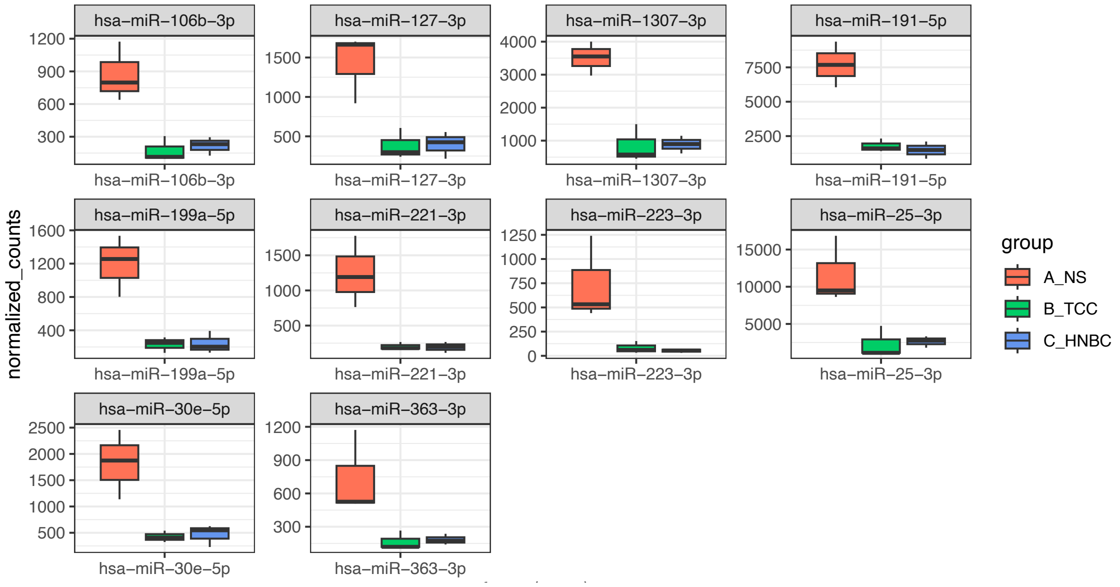


**Supplementary figure S2**

**Count data of the RNAseq.** Boxplot showing the counts obtained for the miRNAs selected for functional analyses. Boxes show the median with range value. Full data are available in the Supplementary Table 1

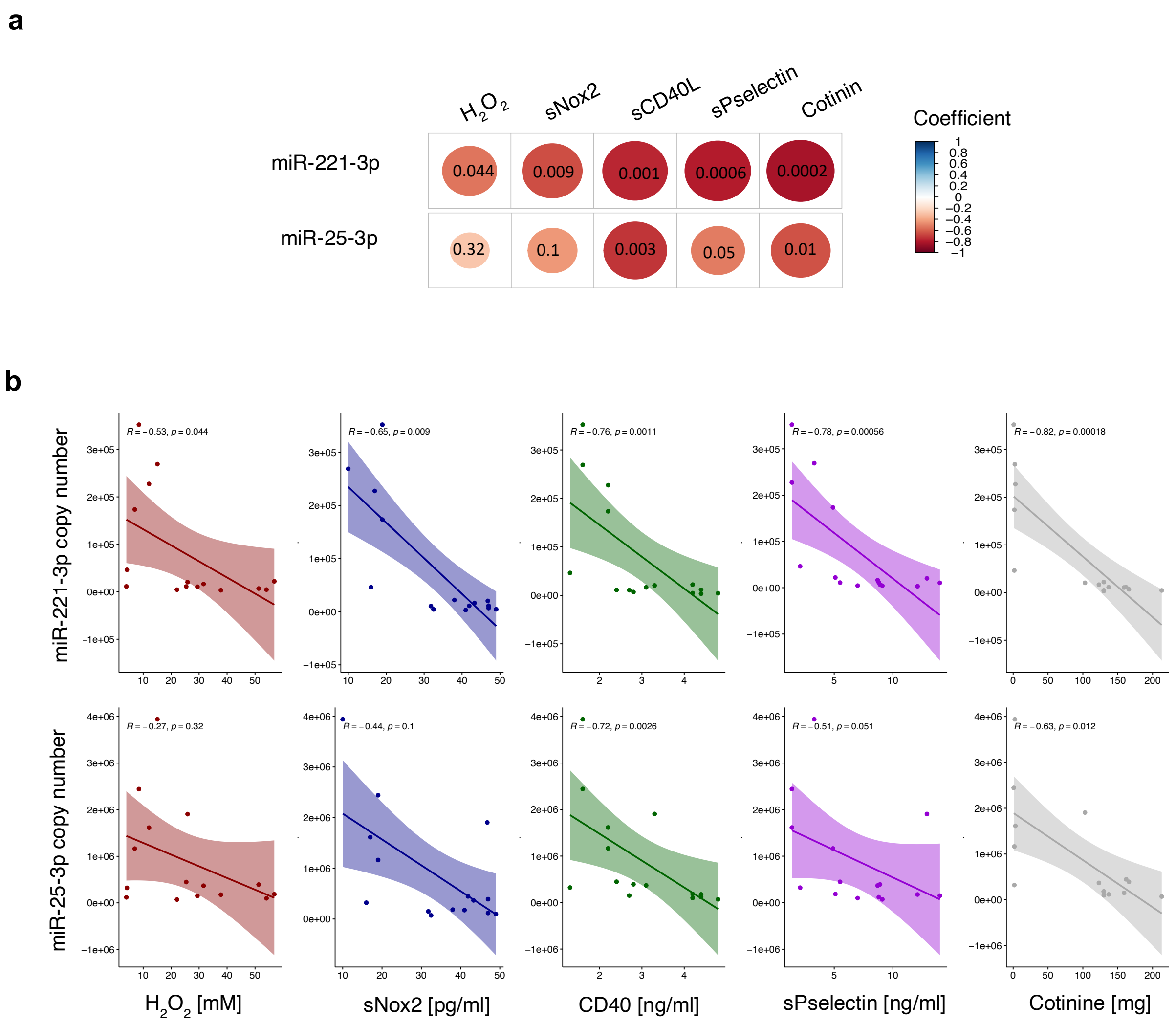


### Supplementary Figure S3

**The abundance of two miRNAs in serum, miR-25-3p and miR-221-3p, show a negative correlation with data relative to oxidative stress and vascular function.** a) Correlogram showing the correlation between the copy number of miR-221-3p or miR-25-3p and the blood parameters measured in the five samples of the validation set. The color shows the correlation coefficient value and the size of the circle is proportional to the extent of correlation. The p value is indicated in each circle. b) panel showing the correlation plots of each variable and the level of each miRNAs in the validation set. The data were analysed using Spearman rank correlation. R coefficient and p value are shown in each graph.
